# An Inherently Interpretable AI model improves Screening Speed and Accuracy for Early Diabetic Retinopathy

**DOI:** 10.1101/2024.06.27.24309574

**Authors:** Kerol Djoumessi, Ziwei Huang, Laura Kühlewein, Annekatrin Rickmann, Natalia Simon, Lisa M. Koch, Philipp Berens

**Affiliations:** Hertie Institute for AI in Brain Health, University of Tübingen, Germany; Tübingen AI Center, University of Tübingen, Tübingen, Germany; University Eye Hospital, University of Tübingen, Tübingen, Germany; Eye Clinic Sulzbach, Knappschaft Hospital Saar, Sulzbach, Germany; Black Forest Eye Clinic, Endingen, Germany; Department of Diabetes, Endocrinology, Nutritional Medicine and Metabolism UDEM, Inselspital, Bern University Hospital, University of Bern, Switzerland

## Abstract

**Background:** Diabetic retinopathy (DR) is a frequent concomitant disease of diabetes, affecting millions worldwide. Screening for this disease based on fundus images has been one of the first successful use cases for modern artificial intelligence in medicine. Current state-of-the-art systems typically use black-box models to make referral decisions, requiring post-hoc methods for AI-human interaction.

**Methods:** In this retrospective reader study, we evaluated an inherently interpretable deep learning model, which explicitly models the local evidence of DR as part of its network architecture, for early DR screening. We trained the network on 34,350 high-quality fundus images from a publicly available dataset and validated its state-of-the-art performance on a large range of ten external datasets. We obtained detailed lesion annotations from ophthalmologists on 65 images to study if the class evidence maps highlight clinically relevant information. Finally, we tested the clinical usefulness of our model in a reader study, where we compared screening for DR without AI support to screening with AI support with and without AI explanations.

**Results:** The inherently interpretable deep learning model obtained an accuracy of .906 [.900-.913] (95%-confidence interval) and an AUC of .904 [.894 – .913] on the internal test set and similar performance on external datasets. High evidence regions directly extracted from the model contained clinically relevant lesions such as microaneurysms or hemorrhages with a high precision of .960 [.941 - .976]. Decision support by the model highlighting high-evidence regions in the image improved screening accuracy for difficult decisions and improved screening speed.

**Interpretation:** Inherently interpretable deep learning models can reach state-of-the-art performance and support screening for early DR by improving human-AI collaboration.

**Funding:** This work was supported by the Hertie Foundation, the German Science Foundation (BE5601/8-1 and the Excellence Cluster 2064 “Machine Learning — New Perspectives for Science”, project number 390727645), the Carl Zeiss Foundation (“Certification and Foundations of Safe Machine Learning Systems in Healthcare”) and International Max Planck Research School for Intelligent Systems.

## 1. Introduction

The global prevalence of diabetes is believed to have reached 10% of the adult population in 2021 with more than 530 million people affected [1] and will likely further increase. Of these patients, more than 20% will develop diabetic retinopathy (DR), a leading cause of blindness among the working-age population [2] and the third leading cause of vision impairment worldwide after age-related macular degeneration and cataract [3, 4]. Although vision loss is one of the most feared complications of diabetes and yearly screening is recommended [5], more than 20% of patients do not take part in regular eye exams, citing timely access and costs as a major hurdle [6]. For these reasons, screening for diabetic retinopathy (DR) has been one of the first successful use cases for artificial intelligence (AI) in medicine [7], promising fast, cost-effective screening even where insufficient clinical personnel is available. Today, multiple AI systems have received regulatory clearance [8] and have been found useful to triage patients not requiring specialist attention and those with vision-threatening DR, potentially contributing to increased screening adherence [9].

Current state-of-the-art models typically use black-box deep learning approaches to make referral decisions. Model-based referral decisions can be explained with heatmaps obtained post-hoc using gradient-based approaches [10, 11, 12]. However, such explanations are not trustworthy, as the produced heatmaps do not reflect the actual decision-making process of the model, and are prone to spurious correlations [13]. Therefore, their results cannot be easily integrated into the clinical decision-making process, as the lack of trustworthy human-interpretable explanation makes it difficult for clinical professionals to validate the AI system’s results [14, 15]. Alternatively, inherently interpretable deep neural networks with specialized architectures designed for transparent reporting could offer trustworthy explanations, potentially leading to improved clinical decisions [14, 16].

We address this issue and validate an inherently interpretable approach for screening for early DR in a retrospective reader study. Our approach uses a deep learning architecture called sparse BagNets [17, 18], which explicitly models the local evidence for the presence of DR as part of its network architecture (figure 1**b**). Most studies so far have considered the task of screening for moderate non-proliferative DR or more advanced stages [7], although even mild non-proliferative diabetic retinopathy (NPDR) is recommended for close monitoring and careful control of hyperglycemia [5, 19]. We reasoned that the benefit of AI-based explanations and decision support would be most clearly visible for this challenging diagnostic task. Trained on a large publicly available dataset, our model shows high specificity and sufficient sensitivity in detecting mild DR across a large array of datasets. We show that the obtained class evidence maps highlight clinically relevant lesions such as microaneurysms or hemorrhages with high precision. In a clinical user study, we show that the system can be effectively used to guide clinical decision-making, leading to 17.5% improvement in diagnostic accuracy for mild DR and overall about ≈ 25% improvement in time.

**Figure 1:**
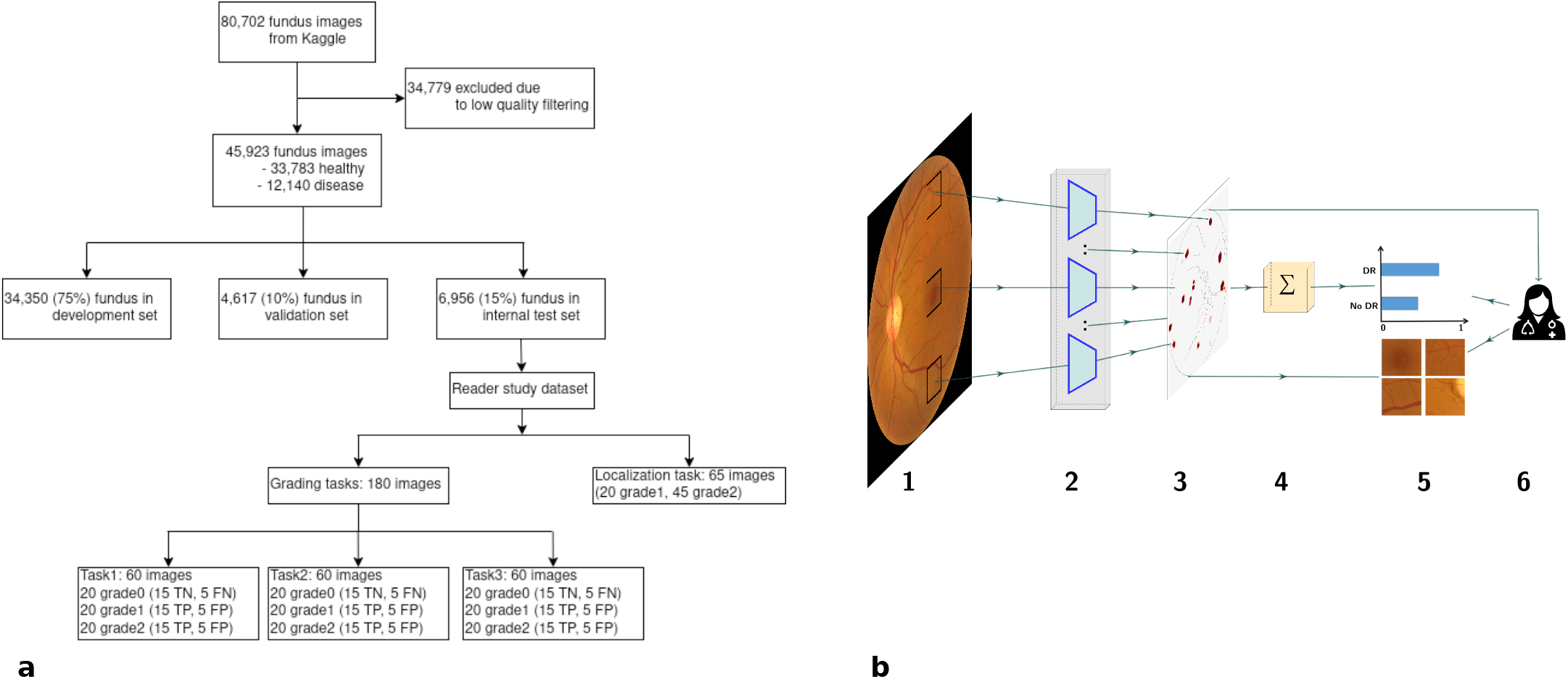
Overview of the development data and proposed inherently interpretable deep learning framework sparse BagNet presented in this study. **(a)** Summary of the development dataset used to build the model, as well as the data used in the retrospective reader study. **(b)** Sparse BagNet architecture. **(b1)** As a preliminary step, the retinal fundus image is implicitly split into many overlapping small patches of size 33 × 33. (**b2**) All patches are fed to the model backbone, which processes them in parallel. (**b3**) The BagNet backbone generates a heatmap that depicts the local disease evidence of individual patches. **(b4)** The values of the heatmap are averaged and used as the final logit for classification. (**b5, b6**) The logits are fed into a softmax function which provides the probability distribution of the output, and then patches of suspect regions based on the heatmaps can be requested and viewed by a clinician to understand the classification results.

### Research in Context

#### Evidence before this study

We searched Pubmed up to 31/05/2024 using the terms “interpretable machine learning” AND “clinical decision support”. We identified 17 articles, of which almost all used features derived from clinical knowledge together with classical machine learning techniques such as logistic regression, decision trees or support vector machines, often combined with post-hoc explainability methods such as Shapley values. One study used deep neural networks with a transformer architecture. None of the studies evaluated the usefulness of their frameworks in a reader study. Searching for “deep learning” and “diabetic retinopathy” resulted in 704 articles. These studies mostly used different variants of blackbox deep neural network architectures for detecting diabetic retinopathy. If the study discussed interpretability, it typically referred to post-hoc methods such as Grad-Cam.

#### Added value of this study

We evaluated an inherently interpretable deep neural network for early diabetic retinopathy detection. We showed that the model can detect early diabetic retinopathy with state-of-the-art accuracy. The class evidence map extracted directly from the model pointed to clinically meaningful lesions in the fundus image. Providing these for clinical decision support reduced screening time and improved grading accuracy for clinically difficult decisions.

#### Implications of all the available evidence

Our findings imply that inherently interpretable deep learning models can perform well in difficult clinically relevant screening tasks. These models provide direct explanations for their decisions as part of their architecture, making them ideal candidates for use in collaborative AI-human settings such as medicine, where trust in AI models is an issue. While we showed their usefulness as part of a retrospective reader study, future research will need to provide additional evidence in prospective, real-world settings.

## 2. Methods

### 2.1. Dataset description and data preparation

We used eleven publicly available retinal image datasets, consisting of color fundus images from various sources, to develop and evaluate an inherently interpretable deep learning model for early DR detection (table 1). For all datasets, fundus images had assigned reference grades based on the International Clinical Diabetic Retinopathy classification scale [31], which provides a grading scheme ranging from 0 (no DR), 1 (mild NPDR), 2 (moderate NPDR), 3 (severe NPDR) to 4 (proliferative DR) according to DR severity. As our goal was to develop an AI system for early DR screening, we combined class level {0} vs {1,2,3,4}. At stage 1, DR is in most cases asymptomatic, and challenging to detect even for experienced ophthalmologists. As all fundus datasets were fully anonymous, no approval from an Ethics Board was needed for this part of the study.

**Table 1:** Summary of the internal and external validation datasets used to evaluate the models. **“Origin”** refers to the country where the data was collected. **“Lesion”** refers to the number of images in the dataset with lesion annotations. The Kaggle dataset (first row, shaded in gray) is the internal dataset used to evaluate the model, while the other datasets were used for external validation to assess the generalization properties of the trained model.

| Dataset | Origin | Number of images |  |  | Lesion |
| --- | --- | --- | --- | --- | --- |
|  |  | All | Healthy | DR |  |
| Kaggle [20] | USA | 6,956 | 5,118 | 1,838 | 65 |
| IDRiD [21] | India | 512 | 168 | 348 | 81 |
| E-Ophtha [22] | France | 434 | 260 | 174 | 174 |
| FGA-DR [23] | UAE | 1,841 | 101 | 1,740 | 1,740 |
| DIARETDB1 [24] | Finland | 89 | 05 | 84 | 84 |
| DDR [25] | China | 12,513 | 6,265 | 6,248 | 755 |
| DR2 [26] | Brazil | 445 | 300 | 145 | - |
| APTOS [27] | USA | 3,662 | 1,805 | 1,857 | - |
| FCM-UNA [28] | Paraguay | 757 | 187 | 570 | - |
| Messidor-1 [29] | France | 1,200 | 546 | 654 | - |
| Messidor-2 [29, 30] | France | 1,744 | 1,017 | 727 | - |

#### Development dataset

The dataset used to develop the inherently interpretable deep learning model was obtained from the Kaggle Diabetic Retinopathy challenge [20] which initially contained records of 44, 351 subjects with 88, 702 retinal fundus images from both eyes (figure 1**a**). After an automated quality filtering using an ensemble of EfficientNet models [32] trained on the ISBI2020 ^1^ challenge dataset, a total of 45, 923 images from 28, 984 subjects were used for training, with 73% of images in the healthy class and 27% in the DR class. The dataset was split into training, validation, and test folds with 75%, 10%, and 15% of images, respectively, making sure that all images from the same subject were allocated to the same fold. The training fold was used for model fitting, the validation fold for model selection and hyperparameter tuning, and the test fold for internal evaluation.

To evaluate the explanations provided by the explainable sparse BagNet model, three ophthalmologists (authors AR, LaK, and NS with 5, 9, and 14 years of experience respectively) marked the location of DR-related lesions on 65 randomly selected fundus images from the test set (20 grade 1 and 45 grade 2) using a custom-written annotation browser interface (appendix figure A1) based on the Python web framework^2^ Django (version 4.2.1) with a secure PostgreSQL database (version 15.3) and a Javascript front-end (appendix figure A1). Annotators were asked to mark “Microaneurysms (MA)”, “Hemorrhages (HE)”, “Exudates (EX)”, “Soft Exudates (SE)” or “Other” for lesions visible on the fundus image. We combined the annotations of all graders into a consensus annotation for each image (appendix table A3). We also assessed the consistency between ophthalmologists’ annotations by calculating the dice between their annotations, showing that identifying DR-related lesions is a difficult task (appendix table A4).

#### External datasets

Additional fundus data sets were obtained from various sources (table 1) and were used for external evaluation of the model to assess the generalization performance. In addition to reference DR grades, some of these external datasets [21, 22, 23, 24, 25] contained pixel-wise annotations for disease-related lesions. We used these additional annotations to evaluate the performance of the interpretable deeplearning model at localizing DR-related lesions.

#### Preprocessing

Raw fundus images were preprocessed by cropping them to a square size of 512 x 512 pixels using a circle fitting method [33]. Then, image intensities were normalized by the mean and standard deviation of the training set. We applied this preprocessing procedure to all the fundus images from all datasets with the same parameters.

### 2.2. Inherently interpretable deep learning model for Diabetic Retinopathy detection

#### Architecture

We trained and evaluated an inherently interpretable deep convolutional neural network (sparse BagNet [17, 18]) for early DR detection. The sparse BagNet is an implicitly patch-based model based on bag-of-local features and aggregates local evidence from interpretable heatmaps to make predictions (figure 1**b**). It takes a two-dimensional fundus image as input (figure 1**b**.1) and outputs a binary prediction, which indicates the absence or presence of DR, together with the confidence as the probability score.

In contrast to other deep learning models, the sparse BagNet architecture is designed to be inherently interpretable, as the input image is implicitly split into many small, overlapping patches (size *q* = 33x33 pixels corresponding to the size of the model’s effective receptive field with stride *s* = 8; figure 1**b**.1), which are independently processed in parallel (figure 1**b**.2) to compute the local evidence for the presence of DR. The patchwise predicted local evidence values are combined into a single class evidence map corresponding to a downsampled version of the input image (figure 1**b**.3), which then is aggregated using average pooling and passed through a softmax function (figure 1**b**.4) to output the probability distribution of DR (figure 1**b**.5).

Crucially, we employ a *ℓ*_1_-penalty on the local evidence predictions to encourage a sparse class evidence map.

After inference, the model can support screening not only with the final prediction but also with the class evidence map (figure 1**b**.3) highlighting the contribution of small local regions to the final prediction. To this end, the evidence map is upsampled to the full image resolution and overlaid on the input image. In contrast to post-hoc gradient-based methods [13], the class evidence map provided by the sparse BagNet is a transparent part of the actual decision-making process and faithfully captures the local evidence. We supplement the class evidence map by extracting patches from regions with high DR evidence (figure 1**b**.5).

#### Training procedure

We trained the model on the training set by minimising the following loss function including the *ℓ*_1_penalty:

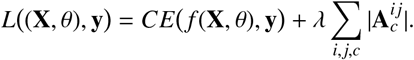

Here, **X** ∈ ℝ^*H*×*W*×*C*^ denotes the input image with *H, W, C* being height, width, and the number of channels, CE is the crossentropy, **y** are the reference class labels, *f* is the model with parameters θ, and **A**_*c*_ denotes the evidence map of class *c*. The sparsity of the evidence maps depends on the hyperparameter λ.

We initialized the model with weights pre-trained on ImageNet and then retrained and optimized for accuracy on the Kaggle DR dataset for 100 epochs (see Sec. 2.1). We used the stochastic gradient descent optimizer with an initial learning rate of 10^−3^, and a clipped cosine learning rate scheduler with a minimum value set to 10^−4^. We performed data augmentation during training by applying random cropping, flipping, color jitter, translation, and rotation following [34]. The sparsity hyperparameter λ was chosen based on the classification accuracy on the validation set (appendix figure A2). For comparison, we trained a standard black-box ResNet-50 [35] using the same settings.

### 2.3. Clinical user study for AI-based decision support

#### Study dataset

The user study was designed to evaluate the usefulness of the explanations provided by the inherently interpretable deep learning model in clinical practice. The dataset for each grading task (see below) consisted of 60 fundus images from the internal test set, where 20 images were sampled from grade 0, grade 1, and grade 2 respectively. For each grade, 15 images were correctly classified by the network and 5 falsely, making this a challenging screening task for clinicians. Thus, the fraction of images with DR in the user study was 66% and the deep learning model achieved an accuracy of 75% by design. Image grading was based solely on the fundus image and AI support, but no additional clinical data were provided.

#### Study design

Six trained ophthalmologists with a median clinical experience of 9 years (4 − 17 years) participated in the reader study (including authors LaK, AR, and NS). The study consisted of three tasks: In task 1 (referred to as “H”), participants were asked to grade fundus images without AI support (appendix figure A3). In task 2 (“H+AI”), participants were additionally provided with the class predicted by the deep learning model and its confidence (appendix A4). Finally, in task 3 (“H+XAI”), participants were additionally shown model explanations in the form of up to 12 bounding boxes around the regions from the class evidence map with the highest evidence, with bounding boxes matching the effective receptive field size and depicting the local image patches that contribute most to the global class evidence (appendix A5).

For the three grading tasks, readers were instructed to classify each fundus image into two classes (“No DR” and “DR”). They were told to classify an image as “DR” even if they thought it only contained signs of mild non-proliferative DR (grade 1). None of the readers had access to the true labels. For task 3, readers were told that some bounding box explanations may contain healthy regions, as the algorithm also generated bounding boxes for healthy images erroneously classified as DR by the sparse BagNet model. In addition to the assigned class, we recorded the time it took for the reader to grade each image and asked them to rate their confidence on a scale from 1 to 5. Ethical approval for the study was obtained from the Ethics Committee at the University Hospital Tübingen (Ref No. 249/2023BO2).

A custom-written browser interface based on the Python web framework Django (version 4.2.1) with a secure PostgreSQL database (version 15.3) and a JavaScript front-end was used to carry out the study (appendix figure A3-A5). The tool showed the fundus image, and response options and provided a digital magnifier to enlarge small image regions.

### 2.4. Evaluation criteria and statistical analysis

Criteria for evaluating the performance of the inherently interpretable deep learning model were specified before the start of the study based on previous work [17]. We evaluated three aspects of the model’s quality:

1. DR screening performance compared to a regular deep learning model, within and across datasets.
2. The quality of the class evidence maps and derived bounding boxes in terms of lesion localization.
3. The usefulness of the inherently interpretable deeplearning model and the derived bounding boxes for decision support.

#### DR screening performance

The primary measure of DR screening performance was the accuracy of the model for early DR detection using the reference labels. Additionally, we evaluated the area under the receiver-operating curve (AUC), sensitivity, specificity, and precision. All measures were computed on the internal test set as well as on the ten external datasets (table 1). The model was not retrained or fine-tuned before assessment on the external datasets. All measures were computed using the scikit-learn package (v 1.0.2) and confidence intervals were computed using a bootstrap procedure with 1000 unstratified resamples [36].

#### Quality of class evidence maps

To measure the quality of the class evidence maps and the derived bounding boxes for lesion localization, we calculated the proportion of highlighted regions (regions within the bounding box) that contained annotated lesions (“localization precision”). To this end, we used the annotations collected for this study on 65 images from the test set, as well as those external datasets containing pixel-level annotations (table 1). We did not evaluate the fraction of lesions detected by our model (“recall”), as we did not train the model for lesion detection, and diagnostic support does not require an exhaustive detection of all lesions.

#### Statistical analysis of decision support

We measured the performance of the readers in our clinical user study (see Sec. 2.3 as the accuracy of the reader’s decision with respect to the reference labels. To assess the effect of the task and DR reference grade statistically, we fit the responses with a generalized linear model (R, function *glm*, v 4.0.3) with predictor *task* or with predictors *task* and *DR grade* including interactions. If we found significant predictors at the *α* = 0.05 level, we computed the marginal means and 95%-confidence intervals (package *emmeans*, v 1.5.3) as well as the respective contrasts between conditions for post-hoc testing. Tukey’s method was used for correcting for multiple comparisons. We used the same procedure for analyzing the measured grading time and the reported confidence, but used a linear model (function *lm*) instead.

### 2.5. Role of the funding source

The funders of this work had no role in the study design, collection, analysis, and interpretation of data, the writing of the report, nor in the decision to submit the paper for publication.

## 3. Results

We trained and evaluated an inherently interpretable deep learning model (“sparse BagNet”) for early DR screening (figure 1**b**, see Sec. 2.2). We first evaluated screening performance for early DR against the state-of-the-art non-interpretable black-box model (“ResNet50”) on the internal test set of the development dataset and a large number of additional datasets (see Sec. 2.1 and table 2).

**Table 2:** Summary of the classification performance with confidence intervals (CIs) computed at 95% using bootstrapping (n=1000) **“AUC”** refer to the receiver-operating curve. **“Loc. precision.”** refers to the localization precision of the sparse BagNet at localizing lesions from annotated images. For each dataset, the first row shows the performance of the interpretable sparse BagNet model, while the second row shows the performance of the baseline black-box ResNet-50 model. The Kaggle dataset (first row, shaded in gray) is the internal dataset used to evaluate the model, while the other datasets were used for external validation to assess the generalization properties of the trained model.

| Dataset | Accuracy | AUC | Sensitivity | Specificity | Precision | Loc. precision |
| --- | --- | --- | --- | --- | --- | --- |
| Kaggle Bag. Res. | .906 (.900 - .913) | .904 (.894 - .913) | .709 (.688 - .729) | .977 (.973 - .981) | .918 (.903 - .932) | .960 (.941 - .976) |
|  | .914 (.907 - .921) | .935 (.927 - .943) | .765 (.745 - .784) | .967 (.962 - .972) | .894 (.878 - .908) | - |
| IDRiD | .891 (.864 - .917) | .879 (.838 - .913) | .951 (.927 - .972) | .768 (.699 - .828) | .895 (.861 - .925) | .811 (.793 - .828) |
|  | .882 (.851 - .909) | .864 (.822 - .902) | .963 (.942 - .981) | .714 (.639 - .781) | .875 (.84 - .908) | - |
| E-Ophtha | .903 (.864 - .917) | .944 (.838 - .913) | .920 (.927 - .972) | .892 (.699 - .828) | .851 (.861 - .925) | .664 (.636 - .692) |
|  | .933 (.851 - .909) | .972 (.822 - .902) | .966 (.942 - .981) | .912 (.639 - .781) | .880 (.840 - .908) | - |
| FGA-DR | .799 (.781 - .819) | .789 (.752 - .823) | .811 (.793 - .830) | .594 (.500 - .687) | .972 (.963 - .980) | .881 (.877 - .886) |
|  | .763 (.743 - .781) | .816 (.768 - .858) | .764 (.743 - .783) | .743 (.653 - .819) | .981 (.973 - .987) | - |
| DIARETDB1 | .831 (.753 - .899) | .931 (.870 - .981) | .821 (.733 - .898) | 1 | 1 | .889 (.870 - .908) |
|  | .742 (.652 - .831) | .811 (.715 - .900) | .738 (.640 - .829) | .800 (.333 - 1.00) | .984 (.950 - 1.00) | - |
| DDR | .825 (.818 - .832) | .926 (.922 - .931) | .669 (.657 - .681) | .980 (.977 - .984) | .971 (.966 - .976) | .965 (.961 - .970) |
|  | .887 (.881 - .892) | .963 (.960 - .966) | .800 (.790 - .810) | .973 (.968 - .977) | .967 (.962 - .972) | - |
| DR2 | .879 (.847 - .908) | .922 (.889 - .951) | .662 (.584 - .742) | .983 (.968 - .997) | .950 (.905 - .990) | - |
|  | .876 (.845 - .906) | .866 (.825 - .905) | .669 (.591 - .742) | .977 (.959 - .993) | .933 (.884 - .975) | - |
| APTOS | .973 (.968 - .979) | .995 (.992 - .996) | .982 (.975 - .987) | .965 (.956 - .973) | .966 (.958 - .974) | - |
|  | .949 (.942 - .956) | .972 (.965 - .978) | .942 (.931 - .952) | .956 (.946 - .965) | .956 (.947 - .966) | - |
| FCM-UNA | .773 (.744 - .802) | .936 (.918 - .952) | .702 (.664 - .738) | .989 (.972 - 1.00) | .995 (.987 - 1.00) | - |
|  | .877 (.853 - .900) | .967 (.954 - .979) | .840 (.811 - .868) | .989 (.971 - 1.00) | .996 (.989 - 1.00) | - |
| Messidor-1 | .889 (.871 - .907) | .943 (.929 - .955) | .832 (.804 - .859) | .958 (.939 - .974) | .959 (.941 - .975) | - |
|  | .893 (.876 - .909) | .954 (.942 - .965) | .852 (.823 - .878) | .943 (.923 - .963) | .947 (.928 - .964) | - |
| Messidor-2 | .829 (.812 - .847) | .876 (.859 - .894) | .750 (.719 - .785) | .886 (.865 - .906) | .825 (.794 - .853) | - |
|  | .851 (.835 - .869) | .925 (.912 - .938) | .794 (.763 - .823) | .893 (.875 - .913) | .841 (.815 - .868) | - |

On the internal test set (table 2, top row), the sparse BagNet performed well and was comparable to the state-of-the-art model (accuracy: 0.906, 95% CI [0.900 − 0.913]; AUC: 0.904 [0.894 − 0.913]; sensitivity: 0.709 [0.688 − 0.729]; specificity: 0.977 [0.973 − 0.981]; precision: 0.918 [0.903 − 0.932]) despite the difficulty of screening for early DR, which includes cases with Non-Proliferative DR (NPDR) with comparably minor abnormalities in the fundus image. Note that these numbers are therefore lower than those reported in other papers for DR screening [7], as most papers evaluate deep learning models for identifying DR starting at moderate NPDR. Despite the difficult task, the inherently interpretable model thus detected 7 out of 10 individuals with a reference label of at least mild NPDR, and the number of false positives was low, with 91 out of 100 positively screened individuals having a DR reference label.

The performance of the sparse BagNet also generalized to external datasets, which partially exhibited strong distribution shifts compared to the development dataset due to the different origin composition (table 1, second column). On most datasets, the model achieved similar performance as on the development dataset, as well as similar performance to the state-of-the-art black-box model. The particularly low performance on the FCM-UNA and FGA-DR datasets could be explained by the relatively low quality of most images in the FCM-UNA dataset and the large intensity variation of the FGA-DR dataset (appendix A6). Taken together, our results show that the inherently interpretable sparse BagNet architecture achieves state-of-the-art performance on a wide variety of datasets.

The key advantage of our inherently interpretable model is that the local disease evidence is explicitly represented in a class evidence map (figure 1**b**.3 and 2**a**-**b**). During training, the class evidence map is encouraged to be sparse, such that the final loss function balances prediction accuracy and an interpretable map. At each location in the map, the color indicates the model output for an individual image patch (see Sec. 2.2). We detected the regions with the highest evidence and placed bounding boxes corresponding to the patch size around these points (figure 2**c**). Although the model was never trained with pixel-level annotations or supervision signals other than the image-level DR reference label, the highlighted regions typically contained DR-related lesions such as microaneurisms, drusen, or hemorrhage with high precision (figure 3).

**Figure 2:**
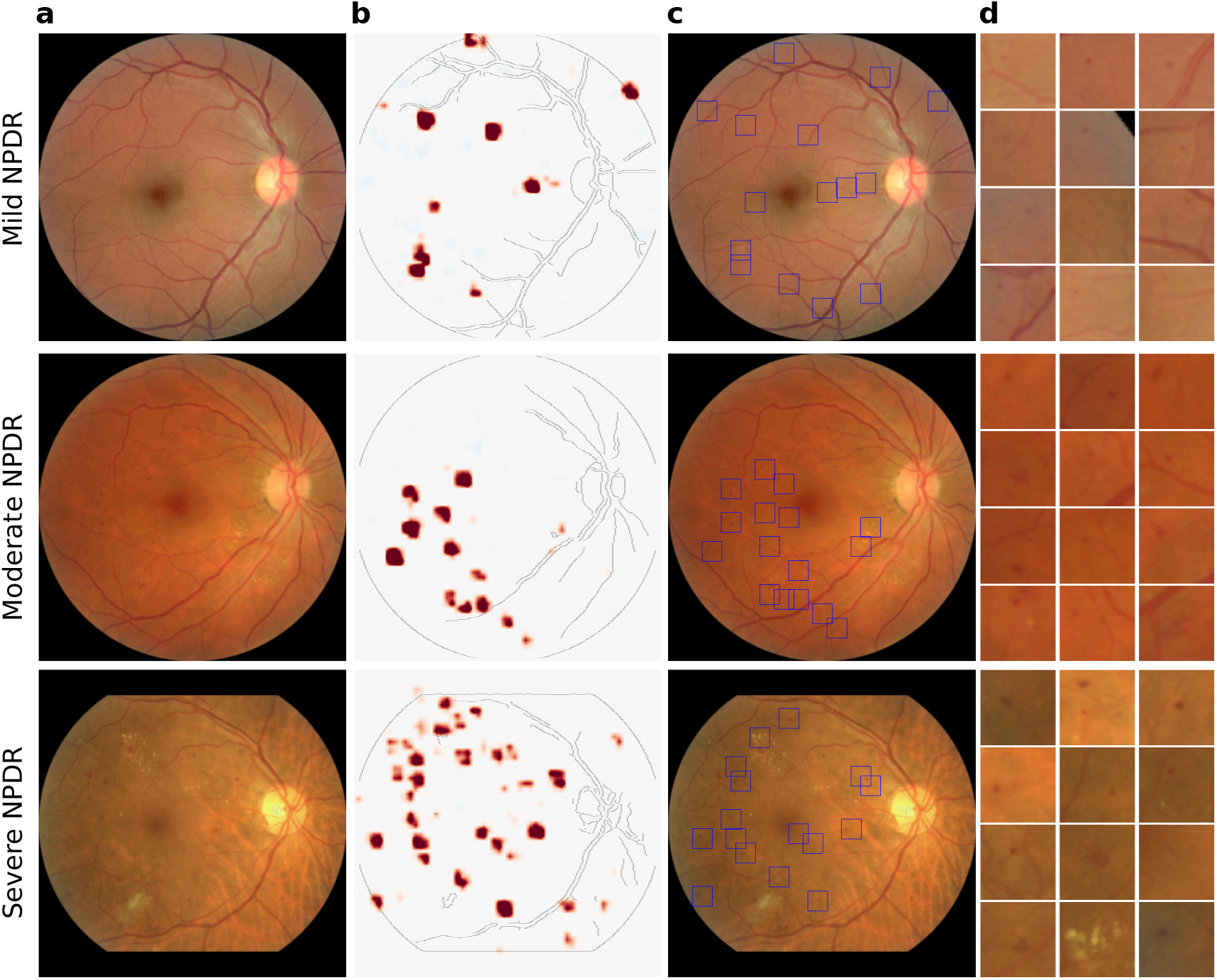
Example images with heatmap visualizations and bounding boxes around relevant regions. **(a)** Examples of retinal fundus images from different DR grades (top to bottom: mild NPDR, moderate NPDR and severe NPDR). **(b)** Heatmap generated by the sparse BagNet, where red regions provide evidence for at least mild DR. **(c)** Bounding boxes around suspicious regions based on the local evidence map. In some cases, the bounding boxes are placed in regions for which there is no visible evidence due to the scaling of the colormap. Yet, these evidence values are also strictly positive. **(d)** Most suspicious regions of **(c)** enlarged and sorted with decreasing evidence scores. Depending on the image grade, the suspicious regions contain various DR-related lesions such as microaneurisms, drusen, or hemorrhage.

**Figure 3:**
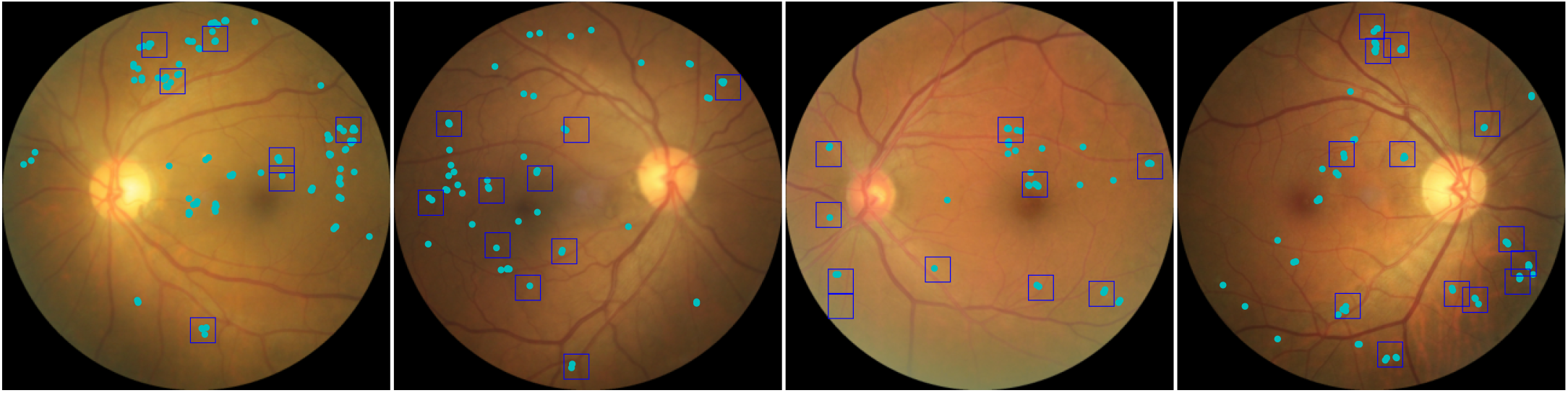
Quality of lesion detection. Example fundus images with DR, with DR lesions (combined annotations by all clinicians) marked as cyan dots. Based on the heatmaps provided by our model, bounding boxes were drawn around the regions with positive local evidence.

We quantitatively evaluated how well the class evidence maps provide information about the location of disease-related lesions using a subset of images from the test set of the development dataset (figure 3) as well as external datasets with pixel-level annotations (table 1). The class evidence maps precisely localized DR lesions, as most regions flagged as suspicious indeed contained annotated lesions (table 2, last column). For the images from the development dataset, we obtained a precision of 0.960 (95% CI [0.941 − 0.976]), with minor differences between images with mild and moderate NPDR (0.783 vs. 0.970). Notably, our model generalized well to external test sets, with precision ranging from 0.664 to 0.965 (table 2, last column). The particularly low localization precision (0.664) on the E-Ophtha dataset could be explained by the fact that annotations were only provided for “Microaneurysms” and “Exudate” lesions, while the images could contain other DR-related lesions. To summarize, the class evidence map extracted from the inherently interpretable sparse BagNet model provided highly precise localization of disease-related lesions.

We then investigated whether our interpretable deep learning model could effectively aid clinicians in detecting DR via a retrospective reader study with six experienced ophthalmologists screening fundus images for the presence of early DR with various levels of AI assistance (see Sec. 2.3).

Without AI assistance (labeled “H”) ophthalmologists reached a mean classification accuracy of 0.611 (95% CI [0.560 − 0.660]; figure 4**a**). Their accuracy increased significantly to 0.758 ([0.711 − 0.800], *p* = 0.0001, post-hoc test with Tukey’s correction for multiple comparisons, see Sec. 2.4) when they had access to the deep learning model’s prediction and confidence (“H+AI”). They achieved similar performance with additional access to AI explanations in the form of bounding boxes around suspicious regions extracted from the class evidence maps (“H+XAI”) at an accuracy of 0.786 [0.741 − 0.825].

**Figure 4:**
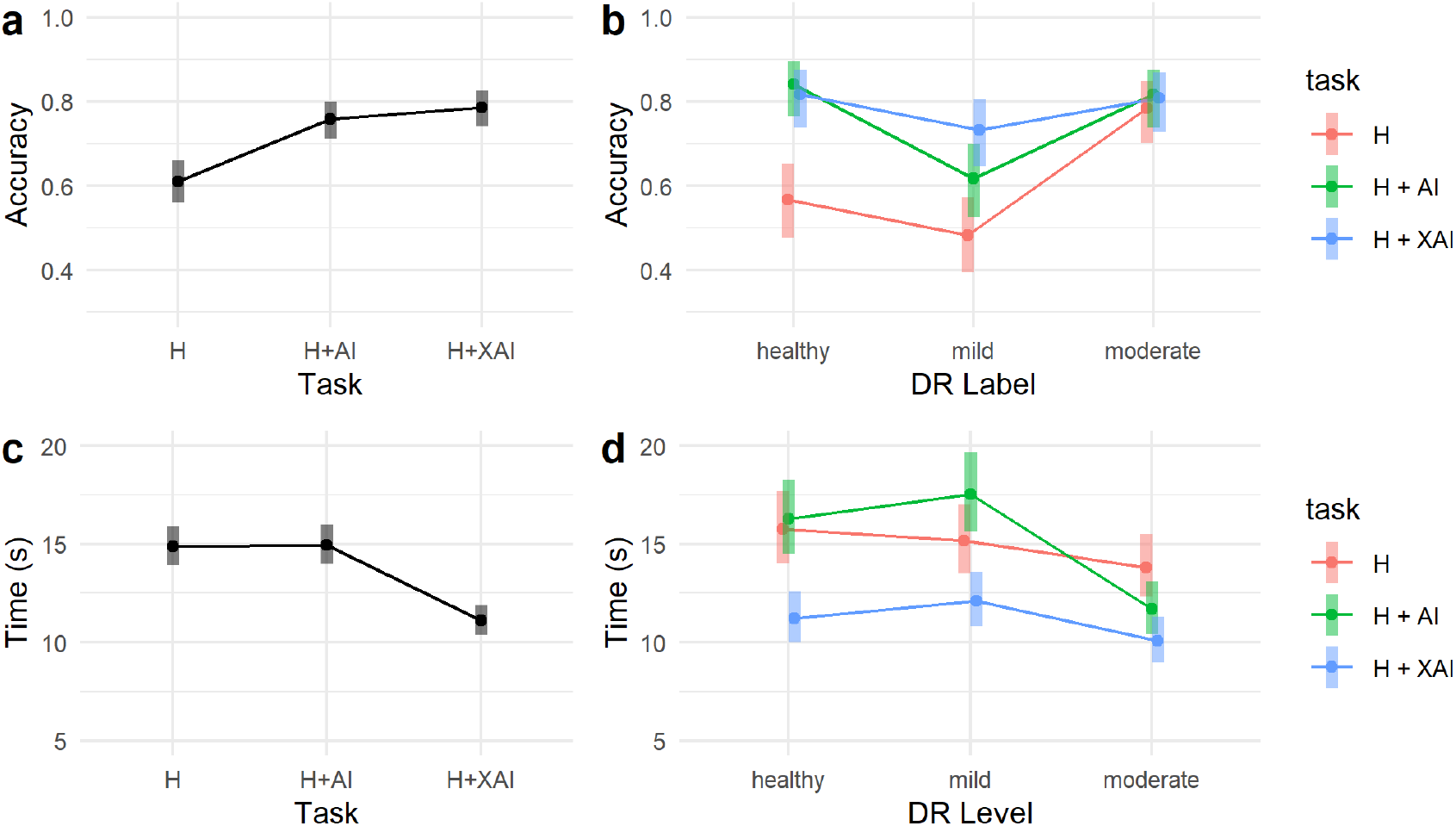
Main results of the retrospective reader study with six experienced ophthalmologists. **(a)** Ophthalmologists’ accuracy with different levels of AI assistance. Ophthalmologists’ accuracy is low without AI assistance “H”, then increases significantly when they have access to AI prediction and confidence “H+AI”, and increases further slightly with additional access to AI explanations “H+XAI”. **(b)** Ophthalmologists’s accuracy in screening for DR on fundus images of different disease grades. For healthy images “grade 0”, accuracy improved significantly with any form of AI decision support (“H+AI” or “H+XAI”) while for images with mild DR (“grade 1”), screening improved significantly for AI support with explanation (“H+XAI”). For images with moderate DR (“grade 2”), AI support had no significant effect on screening performance. **(c)** Ophthalmologists’s decision time in screening DR with different levels of AI assistance. The decision time is significantly reduced with AI support (“H+XAI”) with explanation compared to the other tasks (“H”, and “H+AI”). **(d)** Ophthalmologists’s decision time in screening for DR on fundus images of different disease grades. The reduction in the decision time is present at all disease stages with a significant effect of AI decision support with explanation for healthy images (“grade 0”), mild DR (“grade 1”), and moderate DR (“grade 2”).

We studied ophthalmologists’ performance in screening for DR in fundus images of different disease grades in more detail (figure 4**b**). Without AI support, detecting images with mild DR (grade 1) was the most challenging with comparably low performance, which improved with AI support. For healthy images, screening performance improved significantly with any form of AI decision support (H: 0.567, [0.477−0.652]; H+AI: 0.842, [0.765 − 0.897]; H+XAI: 0.817, [0.737 − 0.876]; H vs. H+AI: *p* < 0.0001; H vs. H+XAI: *p* = 0.0001; H+AI vs. H+XAI: *p* = 0.8645), while for images with mild DR, we observed that screening only improved significantly for AI support with explanations (H: 0.483, [0.395−0.572]; H+AI: 0.617, [0.527 − 0.699]; H+XAI: 0.733, [0.647 − 0.805]; H vs. H+AI: *p* = 0.0962; H vs. H+XAI: *p* = 0.0003; H+AI vs. H+XAI: *p* = 0.1326). For images with moderate DR, AI support had no significant effect on screening performance. Taken together, this provides evidence that giving ophthalmologists access to AI support led to superior DR screening performance, with explanations based on the sparse BagNet model being most effective for difficult diagnostic decisions.

We next studied whether AI decision support would not only allow ophthalmologists to make more accurate screening decisions but also reach their decisions faster. We found that the decision time was significantly reduced when providing ophthalmologists AI support with explanations compared to both other tasks (figure 4**c**, H: 15.2 s [14.1-16.4]; H+AI: 15.9 s [14.7-17.1]; H+XAI: 11.7 s [10.8-12.6]; H vs. H+AI: *p* = 0.7435; H vs. H+XAI: *p* < 0.0001; H+AI vs. H+XAI: *p* < 0.0001). This reduction was present at all disease stages, with a significant effect of AI decision support with explanations for healthy images (figure 4**d**; H: 15.8 s [14.1-17.7]; H+AI: 16.3 s [14.5-18.3]; H+XAI: 11.2 s [10.0-12.6], H vs. H+AI: *p* = 0.9153; H vs. H+XAI: *p* < 0.0001; H+AI vs. H+XAI: *p* < 0.0001), mild DR (H: 15.2 s [13.5-17.0]; H+AI: 17.5 s [15.6-19.7]; H+XAI: 12.1 s [10.8-13.6], H vs. H+AI: *p* = 0.1843; H vs. H+XAI: *p* = 0.180; H+AI vs. H+XAI: *p* < 0.0001), as well as moderate DR (H: 13.8 s [12.3-15.5]; H+AI: 11.7 s [10.4-13.1]; H+XAI: 10.1 s [9.0-11.3]; H vs. H+AI: *p* = 0.1058; H vs. H+XAI: *p* = 0.004; H+AI vs. H+XAI: *p* = 0.1724). In summary, this indicates that decision support with accurate explanations provided by the sparse BagNet model could reduce screening times across all disease levels.

We also analyzed whether AI decision support would change the confidence with which the ophthalmologists could grade the images, but did not find a significant effect of AI support (H: 3.8 [3.7-3.9]; H+AI: 3.7 [3.6-3.9]; H+XAI: 3.6 [3.5-3.7], H vs. H+AI: *p* = 0.6806; H vs. H+XAI: *p* = 0.0543; H+AI vs. H+XAI: *p* = 0.3023). We conclude that self-reported confidence may not be a reliable measure of grader uncertainty compared to recorded decision time.

We finally analyzed whether the positive effect on accuracy was dependent on whether the deep learning model had classified the image correctly or not, as AI support has been reported to be detrimental in case of model errors [37]. In line with the results above, we found that screening performance and decision time significantly improved for cases in which the deep learning model had made a correct decision (appendix figure 4**a**-**b**; accuracy, H vs. H+AI: *p* < 0.0001; H vs. H+XAI: *p* < 0.0001; H+AI vs. H+XAI: *p* < 0.0001; time, H vs. H+AI: *p* = 0.8178; H vs. H+XAI: *p* < 0.0001; H+AI vs. H+XAI: *p* < 0.0001). For cases in which the model had made an incorrect decision, we neither detected positive nor negative effects on accuracy (H vs. H+AI: *p* < 0.3216; H vs. H+XAI: *p* = 0.4953; H+AI vs. H+XAI: *p* = 0.9480) and slightly positive effects on decision time (H vs. H+AI: *p* = 0.4557; H vs. H+XAI: *p* = 0.0941; H+AI vs. H+XAI: *p* = 0.0031).

## 4. Discussion

### Summary of the findings

In this study, we trained and evaluated an inherently interpretable deep learning model for early diabetic retinopathy detection, which is a challenging task even for experienced ophthalmologists. Our model achieved a classification performance comparable to the black-box baseline model in the internal test set and on ten publicly available external datasets. In addition to a binary diagnostic decision, our model provides explanations via interpretable evidence maps, which highlight regions of the image used by the network in making its decisions. In a retrospective reader study, we found that highlighting these regions during grading helped ophthalmologists improve their grading performance, especially for difficult cases, while reducing their decision time. Our study further showed that the errors of the AI model did not negatively affect decision-making by ophthalmologists, in contrast to earlier human-AI studies [37, 38]. A limitation of our model is that it was trained on a dataset from North America, and may need to be fine-tuned on data from the intended target population, although its generalization results on ten additional datasets were promising.

### Need for interpretable AI in medicine

As the potential of AI for medical image analysis has become evident [39, 40], such systems have reached performance close to, or even superior to, those of clinical experts in a variety of tasks [41]. More recently, the focus has shifted towards AI systems assisting clinicians in making better decisions [37]. In this setting, clinicians need to understand how decisions are formed by the AI model, such that transparency and interpretability of medical AI systems have become important aspects [13, 14, 15, 42]. In agreement, the need for trustworthy and transparent AI systems and effective human/AI collaboration has been identified in standardized guidelines to facilitate their adoption in clinical practice [43, 42]. While this generally poses challenges in balancing high performance and interpretability [43], our study has shown that inherent interpretability can be achieved without significant performance trade-offs if the inductive biases of the interpretable model are met – in our case, as early DR causes only very localized lesions in the retina. Such a model can assist clinicians in mitigating the challenge of early and accurate diagnosis of presymptomatic diseases, such as diabetic retinopathy detection. One limitation of our model is that it may not provide good explanations if its inductive bias is not matched to the disease, e.g. when lesions cover large parts of the retina as in more advanced DR grades [17].

### Validation of AI systems in real-world application settings and application readiness

To improve the integration of AI systems in clinical settings, their design must be carried out in collaboration with the identified stakeholders for whom the AI system is intended, to ensure that the resulting model combines different expertise from the beginning and meets the clinical task for which it was developed, including clinicians and patients. Early planning of the intended use of a medical AI model is critical, as it allows to maximize its alignment with the clinical application, reducing the potential for complications at later development stages or during application. Despite the success of AI models in healthcare, comparatively few studies have evaluated their usefulness in collaboration with healthcare professionals in real or retrospective clinical settings [41, 44].

Here, we validated our model for clinical decision support in early DR in a retrospective, simulated clinical reader setting using an online platform. Given the clear evidence for its usefulness in screening for early DR, the next step towards clinical readiness would be to evaluate the system in a prospective study in a dedicated screening setting, e.g. in specialized diabetic clinics, as done for breast cancer screening in [38]. Once the system has also been validated in this context, there is a comparably straightforward path towards deployment as a medical product, as already several similar systems for DR screening have received regulatory clearance [8] and could be upgraded with an interpretable model. We believe that following this route may enable more accurate and faster DR screening, particularly in low-resource settings where the prevalence of diabetes is high and there is a shortage of ophthalmologists to monitor patients’ eye conditions.

## Data Availability

All data produced are available online.

https://github.com/berenslab/retimgtools/releases/tag/v1.1.0

https://github.com/kdjoumessi/Sparse-BagNet_clinical-validation

## 5. Data Sharing

The implementation of our sparse BagNet model is available at GitHub^3^. The annotations performed for this study on selected Kaggle database images, the study data, and the analysis are available in the same GitHub repository.

## 6. Declaration of Interests

The authors declare no competing interests.

## 7 Acknowledgments

We thank Sarah Müller, Pearse Keane, Tunde Peto and Wanjiku Mathenge for discussion.

---

**Figure A1.**
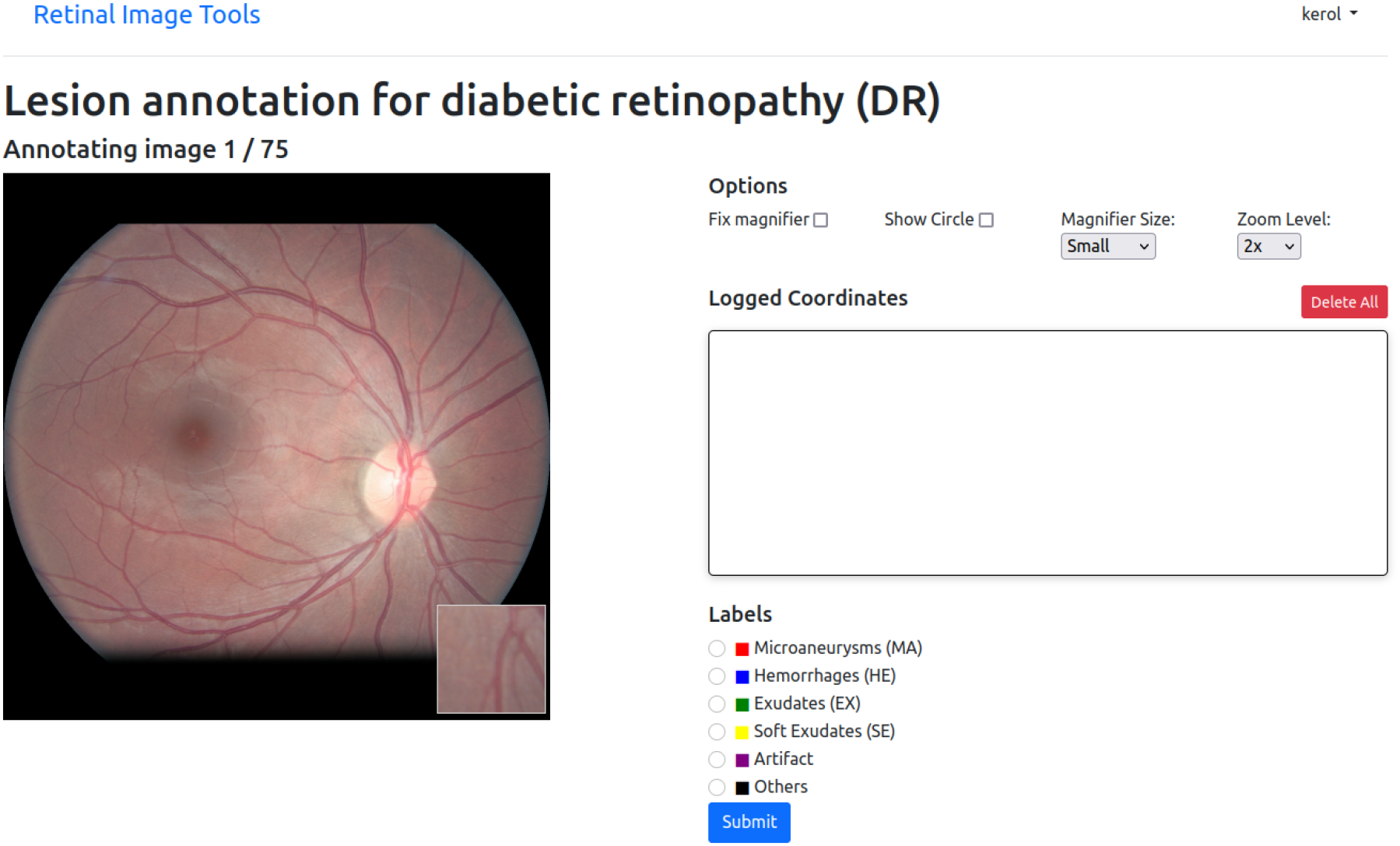
Web interface for the annotation task. A fundus image is shown and based on it, the annotator is asked to annotate lesions related to Diabetic Retinopathy. By moving the mouse over a region of the image, an enlarged version of that region is displayed. All images are from patients with DR of grade 1 (“mild DR”) or 2 (“moderate DR”). Each lesion is marked by selecting the type (Microaneurysms: MA, hemorrhages: HE, exudates: EX, soft exudate: SE, artifact, or any other lesions) and clicking on the image location.

**Table A3.** Summary of model performance on localizing DR-related lesions from graders’ annotations. The precision of the model on each clinician annotation is calculated as the proportion of bounding boxes from regions highlighted on heatmaps containing lesions annotated by a grader. The random precision is obtained by drawing 20 random bounding boxes over each annotated image, excluding those falling in regions containing more than 10% black pixels. The union “∪” gives the precision of the model with the combined clinicians’ annotation masks, while the intersection “∩” gives the precision of the model with reference annotation masks obtained as the intersections of clinicians’ annotation over each image.

|  | Precision |
| --- | --- |
| Grader 1 | 0.709 |
| Grader 2 | 0.610 |
| Grader 3 | 0.923 |
| Random | 0.102 |
| Grader 1 ∪ Grader 2 ∪ Grader 3 | 0.932 |
| Grader 1 ∩ Grader 2 ∩ Grader 3 | 0.545 |

**Table A4.** Inter-grader performance on 65 fundus images from the internal Kaggle test set annotated by three ophthalmologists. “Grader X - Grader Y” refers to the dice score between grader X and grader Y. The Dice score is calculated for each pair of graders as the overlap between their annotation using a patch size of 33 × 33 pixels corresponding to the receptive field of the model and considering different strides (s = 8, 32 for overlapping patches and s=33 for non-overlapping patches). “Grader X - Grader Y ∪ Grader Z” refers to the dice score between grader X, Y, and Z while “Grader Y ∪ Grader Z” is the union between grader Y and Z, and “Grader Y ∩ Grader Z” is the intersection between grader Y and Z.

|  | Dice (s=8) | Dice (s=32) | Dice (s=33) |
| --- | --- | --- | --- |
| Grader 1 - Grader 2 | 0.609 | 0.613 | 0.597 |
| Grader 1 - Grader 3 | 0.545 | 0.543 | 0.542 |
| Grader 2 - Grader 3 | 0.494 | 0.485 | 0.504 |
| Grader 1 - Grader 2 ∪ Grader 3 | 0.546 | 0.551 | 0.544 |
| Grader 2 - Grader 1 ∪ Grader 3 | 0.480 | 0.479 | 0.486 |
| Grader 3 - Grader 1 ∪ Grader 2 | 0.613 | 0.613 | 0.616 |
| Grader 1 - Grader 2 ∩ Grader 3 | 0.609 | 0.602 | 0.597 |
| Grader 2 - Grader 1 ∩ Grader 3 | 0.480 | 0.479 | 0.486 |
| Grader 3 - Grader 1 ∩ Grader 2 | 0.402 | 0.391 | 0.404 |

**Figure A2.**
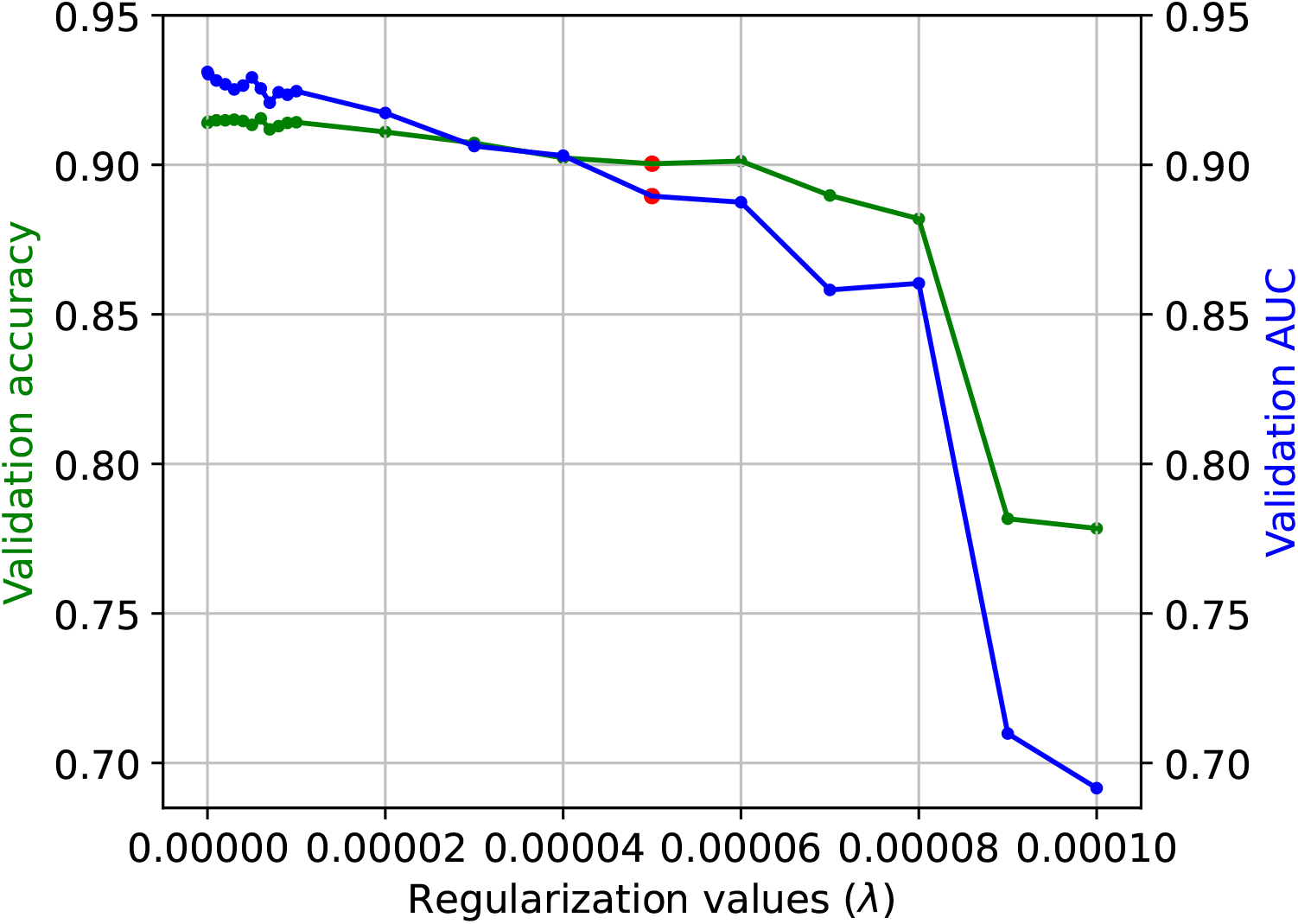
Comparison of the sparse BagNet performance with different regularization values on the validation dataset. The regularization coefficient *λ* affects the classification performance (accuracy and AUC) of the model. The red points indicate the selected value, which is a compromise between sparsity and both accuracy and AUC. It also defines the trade-off between the model’s interpretability and classification performance.

**Figure A3.**
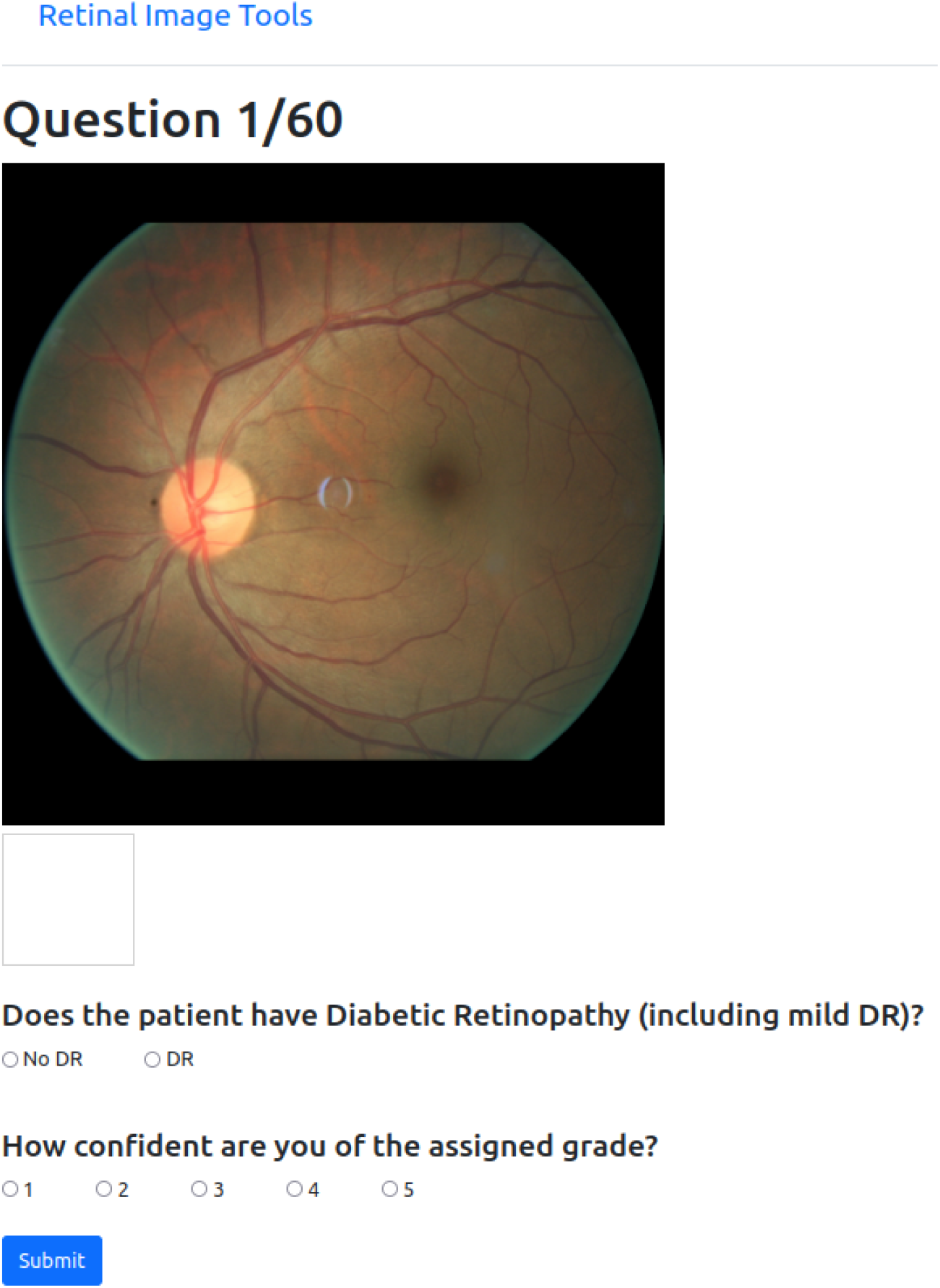
Web interface for the grading task without AI support (“H”) A fundus image is shown and based on it, the grader is asked to decide whether the corresponding patient has Diabetic Retinopathy (DR) of any severity, including mild DR. In addition, the grader is asked to rate the confidence of his/her decision on a scale from 1 (least confident) to 5 (most confident). By moving the mouse over a region of the image, an enlarged version of that region is displayed. The time taken to reach each decision (grading and confidence) is recorded.

**Figure A4.**
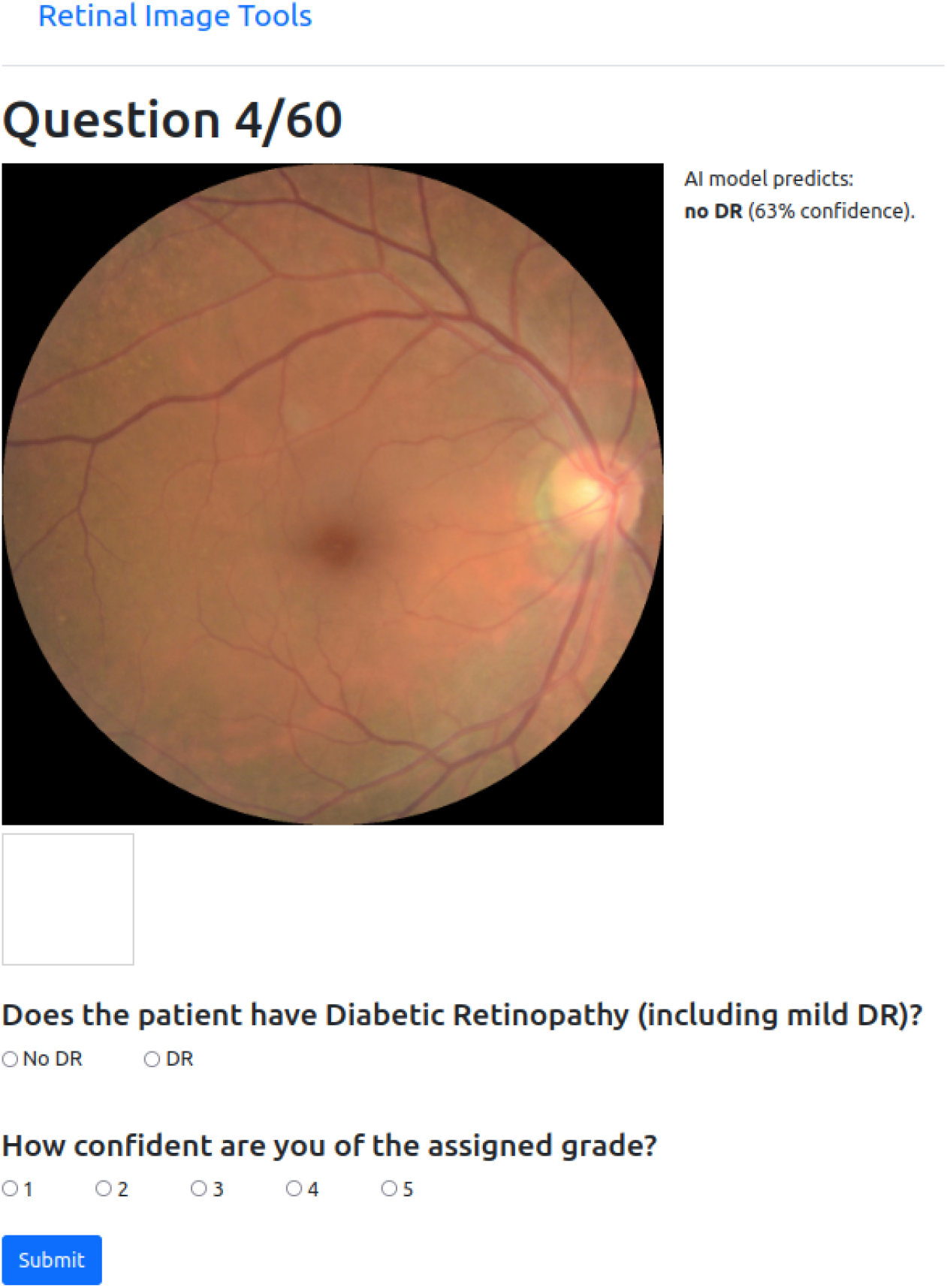
Web interface for the grading task with AI support (“H + AI”) A fundus image is shown with the model’s prediction and its confidence level (from 0% to 100 %, with 100% being the highest confidence score). Based on this, the grader is asked to decide whether the corresponding patient has Diabetic Retinopathy (DR) of any severity, including mild DR. In addition, the grader is asked to rate the confidence of his/her decision on a scale from 1 (least confident) to 5 (most confident). By moving the mouse over a region of the image, an enlarged version of that region is displayed. The time taken to reach each decision (grading and confidence) is recorded.

**Figure A5.**
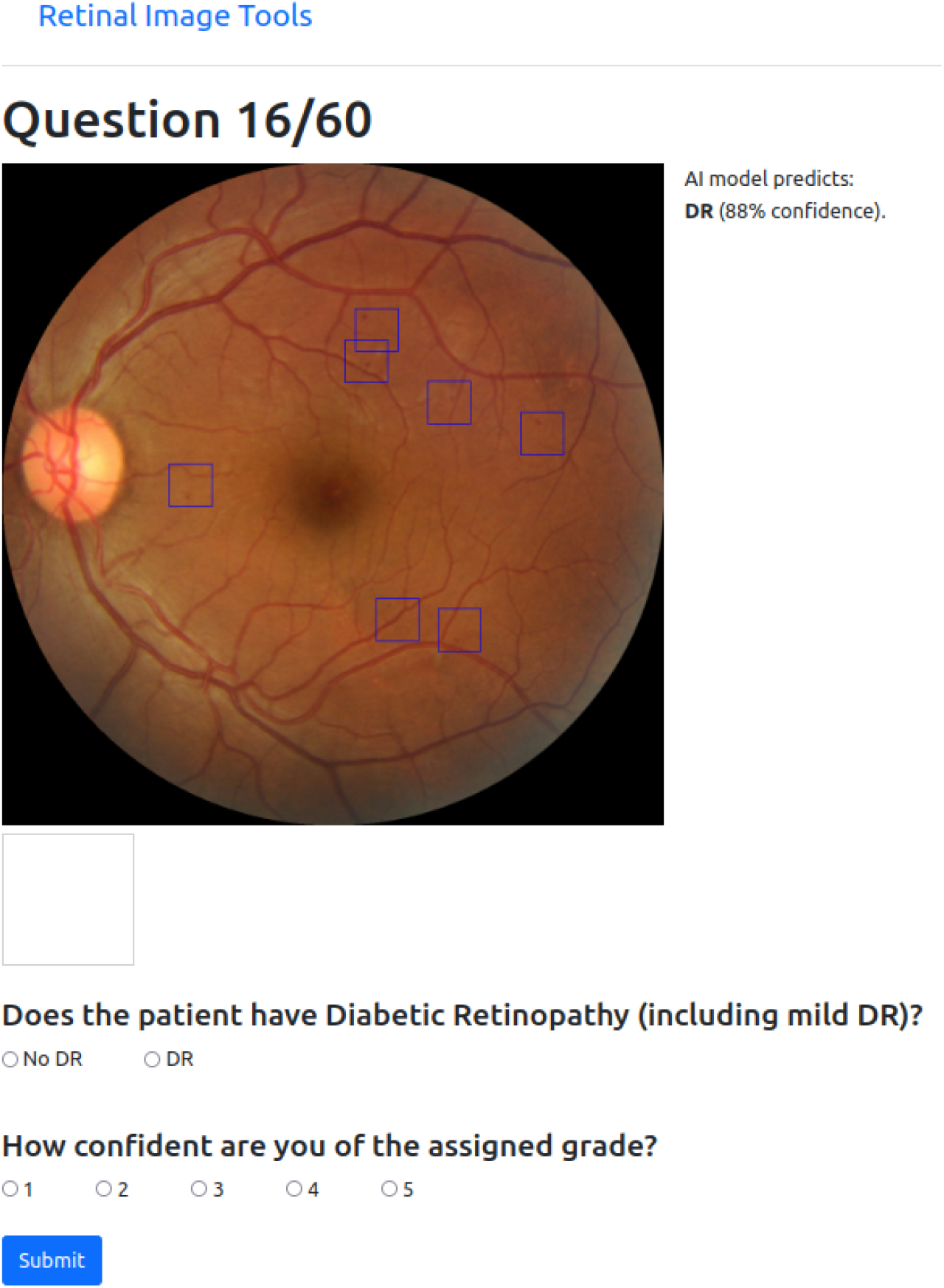
Web interface for the grading task with AI support and explanations (“H + XAI”) A fundus image is shown with the model’s prediction, its confidence level (from 0% to 100 %, with 100% being the highest confidence score), and explanation in the form of blue bounding boxes around the regions for which the AI model believes that they contain signs of DR. Based on this, the grader is asked to decide whether the corresponding patient has Diabetic Retinopathy (DR) of any severity, including mild DR. In addition, the grader is asked to rate the confidence of his/her decision on a scale from 1 (least confident) to 5 (most confident). By moving the mouse over a region of the image, an enlarged version of that region is displayed. The time taken to reach each decision (grading and confidence) is recorded.

**Figure A6.**
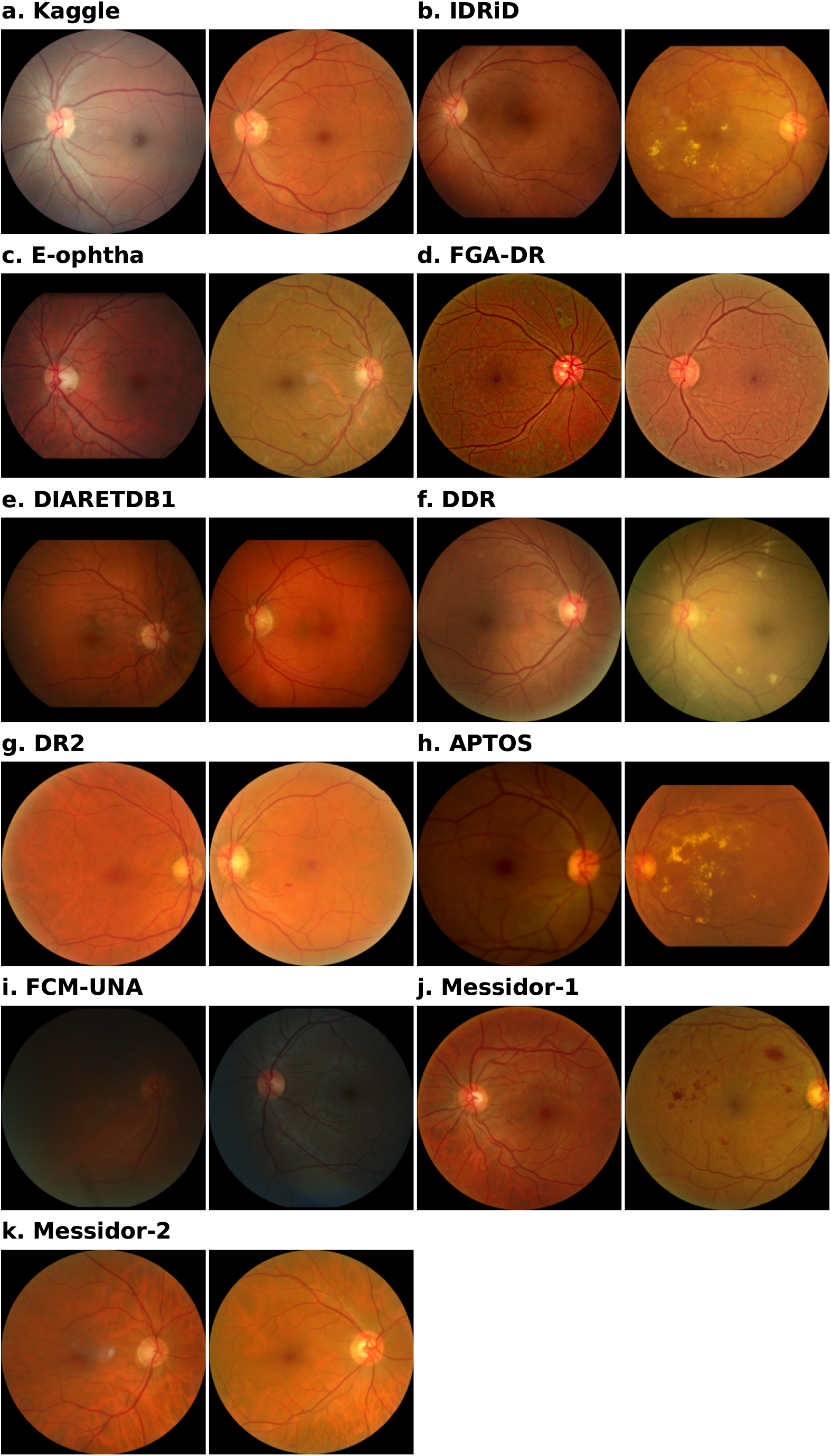
Examples of fundus images from each dataset.

**Figure A7.**
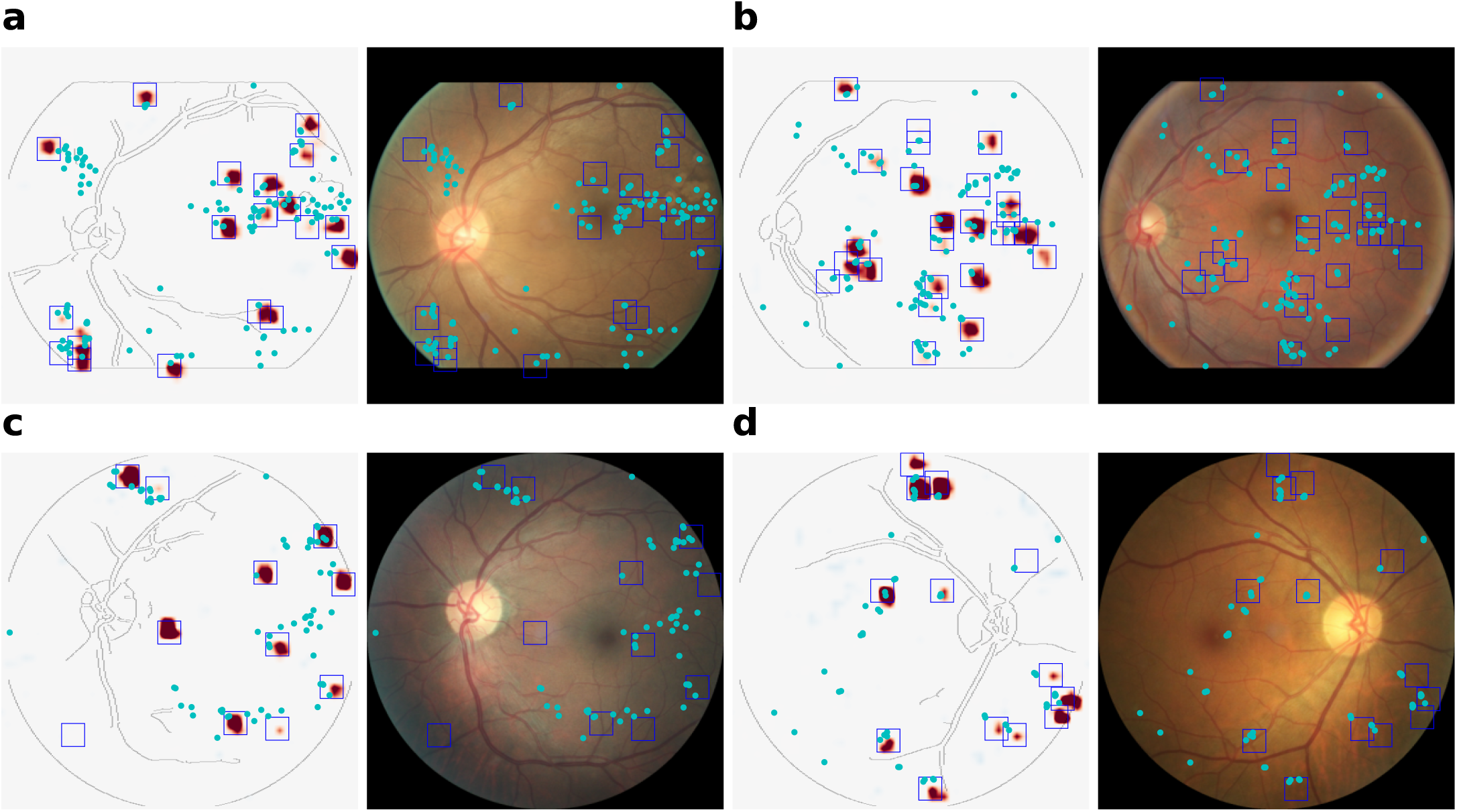
Heatmap with combined clinicians’ annotations of four examples of fundus cases with DR. For each example, the left side shows the heatmap with bounding boxes around the regions of positive activation, while the right side shows the fundus with clinicians’ annotations and bounding boxes around the regions of positive activations. Sometimes, bounding boxes are placed where the positive evidence (in red) is very light and difficult to visualize due to the small number of low positive values.

**Figure A8.**
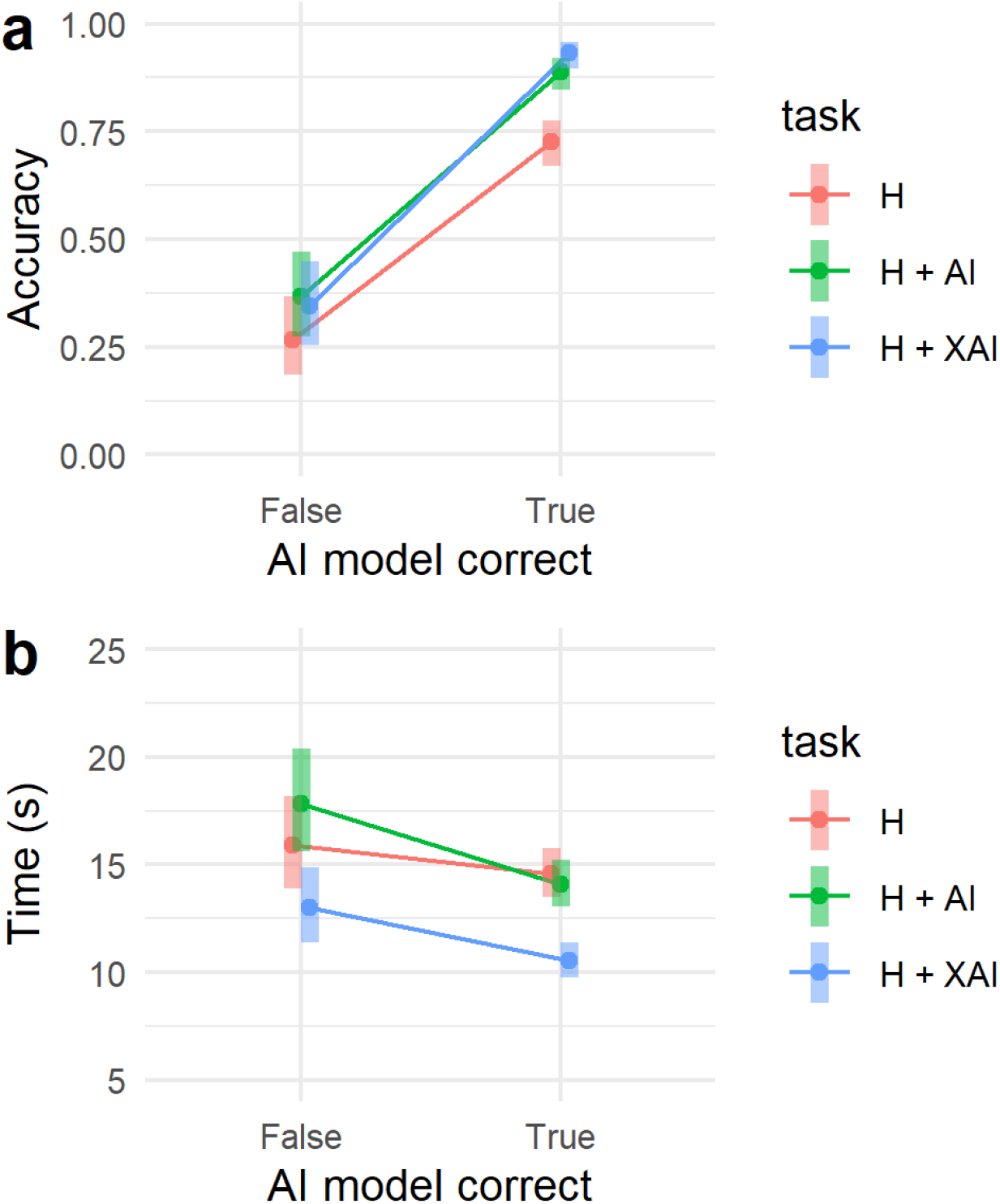
Analysis of errors of the AI model on accuracy and decision times for different tasks during the retrospective reader study. **(a)** For all tasks, ophthalmologists’ accuracy is higher when the deep learning model makes the correct decision. For correct classifications, the AI assistance improves grading accuracy. For incorrect classification, it does not make it worse. **(b)** Ophthalmologists’ decision time decreases overall when the deep learning model makes the correct decision. When the AI model is correct, the explanation decreases decision time significantly, while it does not increase the decision time for incorrect decisions.

## Footnotes

1 https://isbi.deepdr.org/challenge2.html

2 available at https://github.com/berenslab/retimgtools/releases/tag/v1.1.0

3 https://github.com/kdjoumessi/Sparse-BagNet_clinical-validation

## Notes

### Competing Interest Statement

The authors have declared no competing interest.

### Author Declarations

The study used only openly available human data. The main development dataset was originally located at https://kaggle.com/competitions/diabetic-retinopathy-detection. All datasets are described in detail in Table 1 and references 20-29.

